# Neuropsychiatric Sequelae of Brain Injury is Significant, Regardless of Injury Severity

**DOI:** 10.1101/2025.10.08.25337576

**Authors:** Emily S.H. Yeung, Jessica Trier, Elvina M. Chu

**Affiliations:** Brain Injury Rehabilitation Unit, Providence Care Hospital, 752 King St. West, Kingston, Ontario, K7L 4X3

**Keywords:** Brain injury, Neuropsychiatry, Sleep

## Abstract

**Objective:** Brain Injury is the leading cause of death and disability for Canadians under 40 years old. To better understand chronic neuropsychiatric sequelae within our local brain injury population, a retrospective chart review was carried out for all Acquired Brain Injury (ABI) physiatry clinic attendees between November 2014 and December 2021 (n=220) at our public rehabilitation hospital in Southeastern Ontario.

**Methods:** All cases were classified into four subgroups: multiple mild, mild, moderate-severe traumatic brain injury (TBI), and non-TBI. Comparison of depression, anxiety and concussion symptoms were made between subgroups using scores from the Patient Health Questionnaire-9 (PHQ-9), General Anxiety Disorder-7 (GAD-7) and Rivermead Post-Concussion Symptoms Questionnaire, (RPQ), self-reported physical symptoms, sleep disturbance and medications were also investigated. Analysis was repeated with six subgroups, created by further separating cases into moderate and severe TBI where possible.

**Results:** Almost all brain-injury subgroups had moderately severe depression, anxiety and post-concussion symptoms but multiple mild-TBI most frequently self-reported cognitive, neuropsychiatric and sleep disturbance issues. Moderately severe TBI most frequently self-reported physical complaints and sleep disturbance (60%, n=9), although none (0%, n=0) were prescribed sleep medication. Mild TBI (n=60) reported sleep disturbance less frequently (42%, n=25) than moderately severe TBI but 22% (n=13) were prescribed sleep medication.

**Conclusion:** All brain injury subgroups had similar levels of moderately severe neuropsychiatric sequelae; including depression, anxiety and post-concussion symptoms. Multiple mild-TBI had the most self-reported symptoms; while moderately severe TBI were most likely to report physical complaints and sleep disturbance.

## Introduction

Acquired Brain injury (ABI) is the leading cause of death and disability for Canadians under 40 years old (Canada, 2020). Each year, 18,000 people are hospitalized for ABI in Canada (Association, 2012). 6,000 become permanently disabled and 3,000 will be left with physical, cognitive and/or behavioral consequences severe enough to prevent them from returning to pre-injury lifestyles (Agency, 2023). ABI is defined as a non-degenerative and non-congenital brain insult and can be classified as traumatic brain injury (TBI) or non-traumatic brain injury (non-TBI). TBI results from an external mechanical source and is typically clinically classified as mild or moderate-severe (Defrin, 2014), with the majority (80%) being mild. Current TBI literature typically combines moderate and severe subgroups due to difficulties with accurately identifying loss of consciousness (LoC) duration. Non-TBI is due to neuropathology, such as infection or inflammation, leading to brain tissue loss.

Psychiatric symptoms and sleep disturbance are commonly reported following brain injury (King et al., 1995). Changes in cognition, emotion, behaviours, and sleep after ABI can all contribute to neuropsychiatric morbidity, making it particularly challenging to manage clinically (Dikmen et al., 2009). Much research has been focused on identifying neuropsychiatric outcomes following ABI, especially mild TBI as this is the most prevalent subgroup. Guidelines advise that psychopharmacological management is useful, especially following mild TBI (Silver J, 2005) The risk of developing psychiatric disorders including dementia, anxiety, bipolar and psychotic disorders are increased following TBI (Yeh et al., 2020), additionally financial costs due to post-brain injury neuropsychiatric sequelae are 4 times greater for moderate-severe TBI compared to mild TBI (Te Ao et al., 2014). All these factors adversely affect psychosocial functioning post-ABI.

While there is clear evidence showing a link between neuropsychiatric sequelae and TBI, there is limited evidence regarding the symptom profile of various ABI subgroups and whether these may necessitate different treatment approaches. As a result, the best clinical approach for patients with neuropsychiatric sequelae following ABI is unclear and more robust evidence may improve post-ABI neuropsychiatric outcomes.

Instruments reflecting the Diagnostic and Statistical Manual of Mental Disorders (DSM 5-TR) criteria, can be used to screen, diagnose, monitor and measure the severity of neuropsychiatric symptoms of depression and anxiety, post brain-injury (Bell, 1994; Regier et al., 2013). The Patient Health Questionnaire 9 (PHQ-9), is a self-administered, 9-item screening tool, assesses the severity of depressive symptoms and a prospective cohort study has demonstrated that this is also a reliable screening tool for detecting major depressive disorder post-TBI (Fann et al., 2005). The General Anxiety Disorder 7 (GAD-7) questionnaire screens for generalized anxiety disorder (Johnson et al., 2019). The PHQ-9 and GAD-7 have shown high specificity and sensitivity in identifying clinical depression and anxiety disorders in TBI subgroups (Teymoori et al., 2020). Concussion is a traumatically induced transient disturbance of brain function and technically a subset of mild TBI. The Rivermead Post-Concussion Symptoms Questionnaire (RPQ) is a self-report scale measuring post-concussion symptom severity (Potter et al., 2006). Whether used as a self-administered or clinician-administered measure, RPQ scores have been demonstrated to be reliable at assessing symptom severity post-concussion (Potter et al., 2006). The RPQ is however, limited to assessing physical and cognitive symptoms post-injury and does not predict longer-term mental health or measure neuropsychiatric symptoms of depression and anxiety (Asselstine et al., 2020).

The relationship between ABI severity and depression or anxiety symptoms remains largely unknown. A study of 2137 TBI survivors reported similar levels of depression and anxiety across mild, moderate, and severe TBI subgroups; however, authors noted their mild TBI group was 3 times larger than the combined sample of moderate and severe TBI subgroups (Teymoori et al., 2020). While most studies assessing post-ABI depression and anxiety focus on mild TBI, few publications include moderate-severe TBI, or non-TBI subgroups. Sleep disturbance including insomnia, hypersomnia, sleep apnea and narcolepsy are commonly reported post TBI; however, indirect factors such as fatigue, depression, anxiety, medications, and hospitalization arising from TBI also contribute (Aoun et al., 2019).

Post-ABI symptoms may impair ability to return to work. One systematic review found that 50% of individuals with mild TBI did not return to their pre-injury level of function after 1 year (Willemse-van Son et al., 2007) and in those with moderate-severe TBI, 60% were unemployed after 2 years. Another study found little change at 5-years post-injury, where 53% of those with moderate-severe TBI were unable to work (Lo et al., 2021). This suggests brain injury severity may impact functional outcome.

Presenting symptoms and ABI severity should be a guiding factor for clinical decisions regarding prescribing; however, current guidelines lack clarity (Khan et al., 2020). Although the negative impact of post-ABI symptoms is recognized, studies have rarely explored the relationship between ABI severity and neuropsychiatric sequelae (Ponsford et al., 2019).

The primary aim of this retrospective cohort study was to explore and compare the neuropsychiatric sequelae associated with different ABI subgroups. We hypothesize that increased brain injury severity is associated with increased neuropsychiatric symptoms of depression and anxiety, greater sleep disturbance, more self-reported symptom complaints and more frequent use of sleep medication. Return to pre-injury occupation and medications prescribed were investigated as secondary measures of interest.

## Methods

A retrospective cohort study was completed using patient charts from the Physical Medicine and Rehabilitation ABI Clinic. All patients seen in the ABI clinic between November 2014 and December 2021 were initially included (n=229). Nine patients were excluded as 4 charts were missing, 1 had unknown TBI classification, 2 had mixed TBI and ABI, 2 had multiple ABIs. Access to individual electronic and paper medical records was sought through the medical records department after obtaining appropriate ethics approval.

Demographics and outcomes were collected from the most recent clinic attendance including age, sex, date of initial recorded brain injury, date of most recent brain injury clinic attendance, age, self-reported symptoms, ability to continue pre-injury occupation, medication(s) prescribed, plus a score for depression (Patient Health Questionnaire-9, PHQ-9), anxiety (General Anxiety Disorder scale-7, GAD-7), and concussion symptoms (Rivermead Post-Concussion Symptoms Questionnaire, RPQ).

All cases were initially classified into one of 4 subgroups: mild TBI, moderate-severe TBI, multiple mild TBI and non-TBI subgroups. Correlations between subgroups and outcome measures were sought. Secondary analyses of return to work and prescribed medication were included. Analysis was repeated after reclassification into 6 subgroups: mild TBI, moderate TBI, severe TBI, moderate-severe TBI, multiple mild TBI and non-TBI.

Data analyses and figures were completed using Statistical Package for Social Sciences (SPSS) for macOS (SPSS Statistics Premium Campus Edition, version v28.0.1.1, CAN) and GraphPad Prism 9 for MacOS (GraphPad Software Inc., San Diego, CA). Statistical significance was determined by Chi-square test with Bonferroni corrections or one-way ANOVA test with a Tukey’s multiple comparisons unless otherwise stated. *P* value ≤ 0.05 was considered statistically significant.

## Results

A total of 220 patient charts were reviewed, 55% (n=122) men and 45% (n=98) women.

No association was found between ABI subgroup (mild TBI, moderate-severe TBI, multiple mild TBI and non-TBI) and self-rating scale scores for depression (PHQ-9) [F(3, 66)=0.303, p=0.82], anxiety (GAD-7) [F(3, 61)=0.225, p=0.88] or concussion symptoms (RPQ) [F(3, 63)=1.608, p =0.20]. All 4 subgroups had a mean PHQ-9 score of 10 to 14, moderately depressed range and a mean GAD-7 score of 10 to 14, moderately anxious range. The mild TBI, moderate-severe TBI and multiple mild-TBI subgroups had RPQ score >35 indicating moderate to severe concussion symptoms while the severe TBI subgroup appeared less severely affected.

**Table 1.** Four ABI subgroups with corresponding depression, anxiety and concussion symptoms severity shown as mean ± s.d. PHQ-9 score 0-21: 5-9=mild, 10-14=moderate, 15-19=mod-severe depression. GAD-7 score 0-21: 5-9=mild, 10-14=moderate, >15=severe anxiety. RPQ score=0-64: 16-35=post-concussion symptoms affecting function, >35=mod-severe concussion symptoms.

|  | Multiple mild TBI<br>n=7 | Mild TBI<br>n=60 | Mod-Severe TBI<br>n=69 | Non-TBI<br>n=84 |
| --- | --- | --- | --- | --- |
| **PHQ-9** | 12.9±6.7 | 14.0±5.0 | 11.7±7.3 | 11.2±6.9 |
| **GAD-7** | 10.4±6.6 | 12.0±5.6 | 11.6±5.3 | 11.3±6.6 |
| **RPQ** | 38.4±13.9 | 40.6±13.8 | 30.3±12.9 | 48.6±16.4 |

There was a significant difference between ABI subgroups for frequency of self-reported cognitive symptoms X^2^ (3, n=220) =11.83, p=0.008, neuropsychiatric symptoms X^2^ (3, n=220) =9.44, p=.024 and sleep complaints X^2^ (3, n=220) =15.18, p=0.002; with the multiple mild TBI subgroup reporting more symptoms than the mild, moderate-severe, and non-TBI subgroups. Frequency of self-reported physical symptoms was similar for all subgroups X^2^ (3, n=220) =6.03, p=0.11.

There was no relationship between ABI subgroups and return to pre-injury occupation X^2^ (12, n=220) =13.50, p=.334. There was no difference between ABI subgroups and prescription of antidepressants X^2^ (3, n=220) =6.03, p=.108, sleep medication X^2^ (3, n=220) =5.10, p=.165, antipsychotics X^2^ (3, n=220) =2.85, p=.416, benzodiazepines X^2^ (3, n=220) =0.79, p=.852, cannabinoids X^2^ (3, n=220) =0.94, p=.817, mood stabilizers X^2^ (3, n=220) =1.53, p=.675, stimulants X^2^ (3, n=220) =2.12, p=.548 or use of illicit drugs X^2^ (3, n=220) =4.17, p=.244.

Following classification into 6 subgroups to try and better differentiate between moderate and severe TBI, no associations were found between ABI subgroup and depression (PHQ-9) [F(5, 65)=0.677, p=0.64], anxiety (GAD-7) [F(5, 60)=0.745, p=0.59] or concussion symptoms (RPQ) [F(5, 62)=1.515, p=0.20]. Severe TBI appeared to have higher PHQ-9 and GAD-7 scores, indicating worse depression and anxiety symptoms than mild TBI, moderate TBI, non-TBI, moderate to severe TBI and multiple mild TBI subgroups; while moderate TBI reported higher RPQ score, indicating worse concussion symptoms than mild TBI, severe TBI, non-TBI, moderate to severe TBI, and multiple mild TBI subgroups (Table 2) but none of these findings survived Tukey’s multiple comparisons test on ANOVA. In 5 of the 6 subgroups, mean PHQ-9 scores were in the moderately depressed range and in 4 subgroups, mean GAD-7 scores were in the moderately anxious range.

**Table 2.** Six ABI subgroups with corresponding depression, anxiety and concussion symptoms severity shown as mean ± s.d. PHQ-9 score 0-21: 5-9=mild, 10-14=moderate, 15-19=mod-severe depression. GAD-7 score 0-21: 5-9=mild, 10-14=moderate, >15=severe anxiety. RPQ score=0-64: 16-35=post-concussion symptoms affecting function, >35=mod-severe concussion symptoms.

|  | Mild TBI<br>(n = 60) | Mod TBI<br>(n = 15) | Severe TBI<br>(n = 49) | Non-TBI<br>(n = 84) | Mod-severe<br>TBI (n = 5) | Multiple mild<br>TBI (n = 7) |
| --- | --- | --- | --- | --- | --- | --- |
| **PHQ-9** | 12.9±6.7 | 12.5±4.8 | 16.3±4.9 | 11.7±7.3 | 4.0±0.0 | 11.2±6.9 |
| **GAD-7** | 10.4±6.6 | 9.5±3.9 | 15.0±6.0 | 11.6±5.3 | 5.0±0.0 | 11.3±6.6 |
| **RPQ** | 38.4±13.9 | 45.2±10.9 | 27.0±15.6 | 30.3±12.9 | 36.0±0.0 | 41.8±8.2 |

Significant difference was observed in frequency of self-reported physical complaints between subgroups, with moderate TBI having most frequent self-reported physical complaints, this finding survived Bonferroni Correction, X^2^ (5, n=220) =12.02, p=.035. Self-reported cognitive symptoms X^2^ (5, n=220) =3.48, p=.627 and neuropsychiatric symptoms X^2^ (5, n=220) =10.34, p=.066 were similar across all ABI subgroups. Significant difference in self-reported sleep complaints between subgroups also survived Bonferroni Correction X^2^ (5, n=220) =33.10, p<.0001, with moderate TBI (n=15) reporting sleep complaints more frequently (60.0%), although none were prescribed sleep medication. The mild TBI subgroup (n=60) reported sleep complaints less frequently (41.7%) but 22% were prescribed sleep medication.

No relationship between ABI subgroups and return to pre-injury occupation was found X^2^ (5, n=220) =28.79, p=.630. There was no difference between ABI subgroups and prescription of antidepressants X^2^ (5, n=220) =8.95, p=.111, sleep medications X^2^ (5, n=220) =0.18, p=.669, antipsychotics X^2^ (5, n=220) =3.16, p=.676, benzodiazepines X^2^ (5, n=220) =1.53, p=.910, cannabinoids X^2^ (5, n=220) =7.04, p=.218, mood stabilizers X^2^ (5, n=220) =3.26, p=.659, stimulants X^2^ (5, n=220) =3.01, p=.698 or illicit drug use X^2^ (5, n=220) =5.17, p=.396.

## Discussion

In this retrospective chart review study of brain injury, all 4 ABI subgroups (multiple mild TBI, mild TBI, moderate-severe TBI, and non-TBI) had similar moderate to severe levels of depression, anxiety and concussion symptoms. The RPQ scores also suggest moderate to severe limitation in function in all groups except the moderate to severe TBI subgroup who reported being less severely affected by concussion symptoms. The multiple mild TBI subgroup self-reported the most cognitive, neuropsychiatric and sleep problems. Our hypothesis that greater brain injury severity is associated with increased neuropsychiatric symptoms of depression, anxiety and more concussion symptoms was not supported by the findings from our study.

Stratification into 6 ABI subgroups (multiple mild TBI, mild TBI, moderate TBI, severe TBI, moderate-severe TBI, and non-TBI) allowed for greater differentiation between the moderate and severe TBI cases. All subgroups had similar levels of anxiety, depression and concussion symptoms; confirming that greater brain injury severity is not necessarily associated with increased neuropsychiatric sequelae. Self-report of cognitive, neuropsychiatric and sleep issues were most frequent in multiple mild TBI, while moderate TBI had increased self-reported physical symptoms and sleep issues.

The time elapsed between brain injury and clinic appointment when the data was collected varied and could be a confounding factor in this study. Caution is necessary in data interpretation as some subgroups were very small. Only 3 individuals from the severe TBI subgroup had PHQ-9 scores available and of the total sample (n=220), only 73 PHQ-9, 69 GAD-7 and 69 RPQ datasets were available. A similar study exploring the relationship between injury severity and depressive symptoms using the PHQ-9 described mild TBI reporting more depressive symptoms than moderate or severe subgroups (Powell et al., 2019).

The moderately severe TBI subgroup most frequently self-reported physical symptoms and had sleep complaints. A large cohort study of US veterans described an increased risk of incident sleep disorders following TBI, although a stronger association with mild rather than moderate TBI was described (Leng et al., 2021). Our findings suggest that individuals with moderately severe TBI may have more symptoms adversely affecting both physical wellbeing and sleep.

Return to previous occupation was similar between all ABI subgroups. This suggests that even a mild TBI can prevent return to previous occupation and does not necessarily correlate with milder functional impairment, especially considering that our severe TBI and non-TBI subgroups included more people who were unemployed, retired or in formal education prior to their brain injury. In a study by van der Horn et al. (van der Horn et al., 2013) the mild TBI subgroup were more anxious and depressed, therefore more likely to have incomplete return to work. This may help explain our findings as levels of depression and anxiety were moderately severe across all subgroups and return to previous occupation was also similar, regardless of brain injury severity.

Prescription trends for sleep and psychotropic medication were similar across subgroups. Although moderate TBI most frequently reported sleep disturbance, none were prescribed hypnotics. The mild TBI subgroup reported sleep disturbance less frequently, but this may be explained because 1 in 5 were being prescribed hypnotics. Our findings suggest sleep is a prominent issue that should be addressed following any brain injury but those in the moderate TBI category may be at increased risk of having an untreated sleep issue.

The number of people in this study using cannabinoids (n=12) was 3 times greater than the number of people using illicit substances (n=4) post brain injury. Cannabis was legalised in Canada in 2018, yet little is known about the adverse neuropsychiatric effects associated with the ABI population. Cannabinoids have been shown to negatively impact memory, attention, and psychomotor function; these overlap considerably with commonly reported post-TBI symptoms and may risk symptom exacerbation (Hergert et al., 2021). Further research is necessary to fully understand the effect of cannabinoids within the ABI context.

Our study found that brain injury severity is not related to depression, anxiety or concussion symptom severity but these neuropsychiatric sequelae are present at a significant level of symptom severity in all categories of brain injury. Multiple mild TBI was associated with increased self-reporting of cognitive, neuropsychiatric and sleep issues, while moderate TBI was associated with increased self-reporting of sleep disturbance and physical symptoms after the moderate to severe subgroup was further separated out. Functional outcome as measured by return to previous occupation was not related to brain injury severity and medication prescribing was also similar.

In conclusion, neuropsychiatric sequelae of depression, anxiety and post-concussion symptoms are common and severe enough to be clinically relevant following any severity of acquired brain. Differentiating between moderate and severe brain injury may have clinical value in guiding better symptom management and treatment options, allowing characterisation of distinct clinical features such as sleep disturbance. Ready access to psychiatry services within a brain injury clinic setting should help improve patient outcomes.

## Acknowledgements

We are grateful to all at Providence Care Hospital for supporting the work to go ahead.

## Author contributions

J.T. and E.M.C. conceived the study, E.S.H.Y. performed the data collection, E.S.H.Y and E.M.C. analyzed the data, E.S.H.Y. and E.M.C. drafted and revised the paper; all authors approved the final version of the manuscript.

## Data availability statement

All patient medical data collection adhered to the guidelines of the Health Sciences and Affiliated Teaching Hospitals Research Ethics Board (HSREB).

